# Identification of a novel mosaic *MTOR* variant in purified neuronal DNA from depth electrodes in a patient with focal cortical dysplasia

**DOI:** 10.1101/2024.01.18.24301006

**Authors:** Karl Martin Klein, Rumika Mascarenhas, Daria Merrikh, Maryam Khanbabaei, Tatiana Maroilley, Navprabhjot Kaur, Yiping Liu, Tyler Soule, Minette Manalo, Goichiro Tamura, Julia Jacobs, Walter Hader, Gerald Pfeffer, Maja Tarailo-Graovac

## Abstract

**Background:** Recent studies have identified brain somatic variants as a cause of focal epilepsy. These studies relied on resected tissue from epilepsy surgery which is not available in most patients. The use of trace tissue adherent to depth electrodes used for stereo electroencephalography (stereo EEG) has been proposed as an alternative but is hampered by the low cell quality and contamination by non-brain cells. Here, we use our improved depth electrode harvesting technique that purifies neuronal nuclei to achieve molecular diagnosis in a patient with focal cortical dysplasia (FCD).

**Methods:** Depth electrode tips were collected, pooled by brain region and seizure onset zone, nuclei isolated and sorted using fluorescence-activated nuclei sorting (FANS). Somatic DNA was amplified from neuronal and astrocyte nuclei using primary template amplification followed by exome sequencing of neuronal DNA from the affected pool, unaffected pool, and saliva. The identified variant was validated using droplet digital PCR.

**Results:** An adolescent male with drug-resistant genetic-structural epilepsy due to left anterior insula FCD had daily focal aware seizures. Stereo EEG confirmed seizure onset in the left anterior insula. The two anterior insula electrodes were combined as the affected pool and three frontal electrodes as the unaffected pool. FANS isolated 140 neuronal nuclei from the affected and 245 neuronal nuclei from the unaffected pool. A novel somatic missense *MTOR* variant (p.Leu489Met, CADD score 23.7) was identified in the affected neuronal sample. Droplet digital PCR confirmed a mosaic gradient (VAF 0.78% in affected neuronal sample, variant was absent in all other samples).

**Conclusions:** Our finding confirms that harvesting neuronal DNA from depth electrodes followed by molecular analysis to identify brain somatic variants is feasible. Our novel method represents a significant improvement compared to the previous method by focusing the analysis on high quality cells of the cell type of interest.

## Introduction

Recent studies in resected tissue from epilepsy surgery have identified brain somatic variants as a cause of focal epilepsy. These somatic variants are not inherited but occur during brain development and are therefore only present in a subset of cells, often only in the brain (i.e., mosaic). Pathogenic variants that cause overactivity of the mTOR pathway have been identified in focal cortical dysplasia (FCD) type 2 and hemimegalencephaly^1–12^. Pathogenic variants in *SLC35A2* were found in mild malformation of cortical development with oligodendroglial hyperplasia^13^. Genes of the Ras/Raf/MAPK signaling pathway play a role in mesial temporal lobe epilepsy with hippocampal sclerosis^14^ and low-grade epilepsy associated tumors^12^.

The identification of brain somatic variants requires access to brain tissue which was classically provided by resective epilepsy surgery. However, not all patients are amenable to surgery and even in patients undergoing surgery, minimally invasive approaches (such as laser interstitial thermal therapy, LITT) are increasingly being used. The latter destroy the epileptogenic tissue *in situ* rather than remove it. Consequently, novel approaches to identify brain somatic variants are needed in order to allow a molecular diagnosis in a larger group of patients^15^.

One such approach uses trace tissue adherent to depth electrodes used for stereo EEG (sEEG) recordings in epilepsy patients undergoing invasive presurgical workup. This method was first proposed by Montier et al. resulting in the identification of *MEN1* as a possible candidate gene for epilepsy^16^. Electrodes were collected after extraction in phosphate-buffered saline (PBS), followed by pelleting of cells via centrifugation and whole genome amplification of the DNA. The latter increases the minute amounts of DNA to allow comprehensive molecular analysis, for example with exome sequencing. Ye et al. used the same method to identify a mosaic gradient for a *KCNT1* variant in a patient with nonlesional multifocal epilepsy^17^. Three further studies compared variant allele frequencies (VAF) of variants identified in resected tissue samples with the VAF in depth electrode samples from the same patients^18–20^. Across these studies, variants could be identified in depth electrode samples in four out of five patients. VAFs in these four patients were 7-27x lower than in tissue. One explanation for the lower allele frequencies is that only a part of the depth electrode may be located within the area of interest resulting in a dilution of the sample with healthy cells from other areas^18,20^.

However, in our experience there is also significant contamination of the brain cells adhering to the electrodes, for example with immune cells reacting to the implanted electrodes and blood. These contaminating non-brain cells significantly reduce the relative fraction of brain cells that carry the variant which in turn results in lower VAFs. In the worst case, this may result in the inability to identify the underling somatic genetic cause.

To overcome this issue, we have developed a novel depth electrode harvesting technique that uses fluorescence-activated nuclei sorting (FANS) to purify neuronal nuclei and exclude any contaminating cells. We then amplify DNA from purified neuronal nuclei which significantly increases VAFs and the ability to identify the underlying somatic variant. In addition, we are using a recently developed amplification method (primary template amplification, PTA)^20–22^. PTA only amplifies from the primary template and avoids exponential amplification of artifacts associated with multiple displacement amplification that was used by previous studies^16,17^. Here, we use our novel technique to identify a mosaic *MTOR* variant in a patient with left anterior insula FCD.

## Methods

After obtaining written informed consent, a saliva sample was collected, and DNA extracted (DNA Genotek prepIT L2P). Depth electrode (Ad-Tech SD, 5 mm contact spacing) tips were collected in PBS immediately after their removal and frozen at -80°C. Electrodes were pooled by brain region and seizure onset zone.

On the day of processing, samples were thawed on ice and nuclei isolated using the Minute^™^ Single Nucleus Isolation Kit for Neuronal Tissues/Cells (BN-020, Invent Biotechnologies Inc). Nuclei were stained with the conjugated antibodies anti-NeuN (ab190195, Abcam), anti-LHX2 (129079-ML650, USBiological) and anti-SPI1 (658010, Biolegend) as well as DAPI^23^. Nuclei were sorted using FANS into neuronal (DAPI+, NeuN+, SPI1-), astrocyte (DAPI+, LHX+, NeuN-, SPI1-), microglia (DAPI+, SPI1+, NeuN-) and other nuclei (DAPI+, NeuN-, LHX2-, SPI1-). Somatic DNA was amplified separately from neuronal and astrocyte nuclei from each electrode pool (i.e., region) using PTA (ResolveDNA® Whole Genome Amplification Kit 100136, Bead Purification Kit 100182, BioSkryb Genomics). DNA was quantified with the Qubit dsDNA HS kit. Amplified somatic and saliva DNA aliquots were analyzed using a multiplex assay including multiple short tandem repeat loci (STR) for quality control (ABI AmpFlSTR Identifiler Plus PCR Amplification Kit A26364).

Exome sequencing of DNA from the affected and unaffected neuronal sample as well as saliva was performed at the Centre for Health Genomics and Informatics, University of Calgary, on the NovaSeq 6000 platform (Illumina Inc.) after exome capture using the IDT xGen Human Exome V2 panel. FastQC^24^ and Trimmomatic^25^ were used for pre-alignment quality control and trimming of low-quality bases, respectively, followed by alignment with BWA-MEM^26^ to the hg19 reference genome. Somatic variants were identified using Mutect2^27^ in the affected neuronal sample using the unaffected neuronal sample and saliva as control. Identified variants were annotated using Annovar^28^, VCFanno^29^ and SnpEff^30^ and filtered using population databases (gnomAD^31^, TOPmed^32^) to exclude the variants with frequencies larger than 1% in an unaffected population. Additionally, an in-house database was employed to exclude sequencing errors. Filtered variants were manually reviewed for quality using the Integrative Genomic Viewer (IGV)^33^ for visualization.

Droplet digital PCR (ddPCR) was performed for variant validation using a QX200 Droplet Generator (Bio-Rad), QX200 Droplet Reader (Bio-Rad) and a custom TaqMan® SNP genotyping Assay (Thermo Fisher Scientific, forward primer: TGCCACAGTCTTCACTTGCAT, reverse primer: GCCTGGCCCCATTGC, wildtype reporter sequence: TCGAGCCAGCATGCT, variant reporter sequence: TCGAGCCATCATGCT, annealing temperature 58°C) in 60 ng of neuronal and astrocyte DNA from all electrode pools and in saliva. The data was analyzed with the QX Manager Software 2.1 Standard Edition (Bio-Rad). The detection limit of the assay was established using wildtype DNA spiked with decreasing concentrations of mutant gBlock (Integrated DNA Technologies, Inc.).

## Results

We report an adolescent male with drug-resistant genetic-structural epilepsy due to left anterior insula FCD (Fig. 1 A-B) who had seizure onset during the first years of life. He had daily focal aware seizures with abdominal pain and fear associated with body stiffening lasting 1-2 minutes. He would often scream in pain during the seizures. MRI showed FCD in the left most anterior short insula gyrus possibly blending towards the left orbitofrontal and left opercular areas (Fig. 1 A-B). The posterior insular cortex appeared to be normal. Noninvasive video EEG monitoring had shown bilateral frontal and central interictal epileptiform discharges and left frontocentral ictal onset. A subtraction ictal SPECT showed left frontal hyperperfusion. Neuropsychological testing revealed low average performance with weaknesses in executive dysfunction. Clinical genetic testing including whole exome sequencing and chromosomal microarray from peripheral DNA were negative.

**Figure 1:**
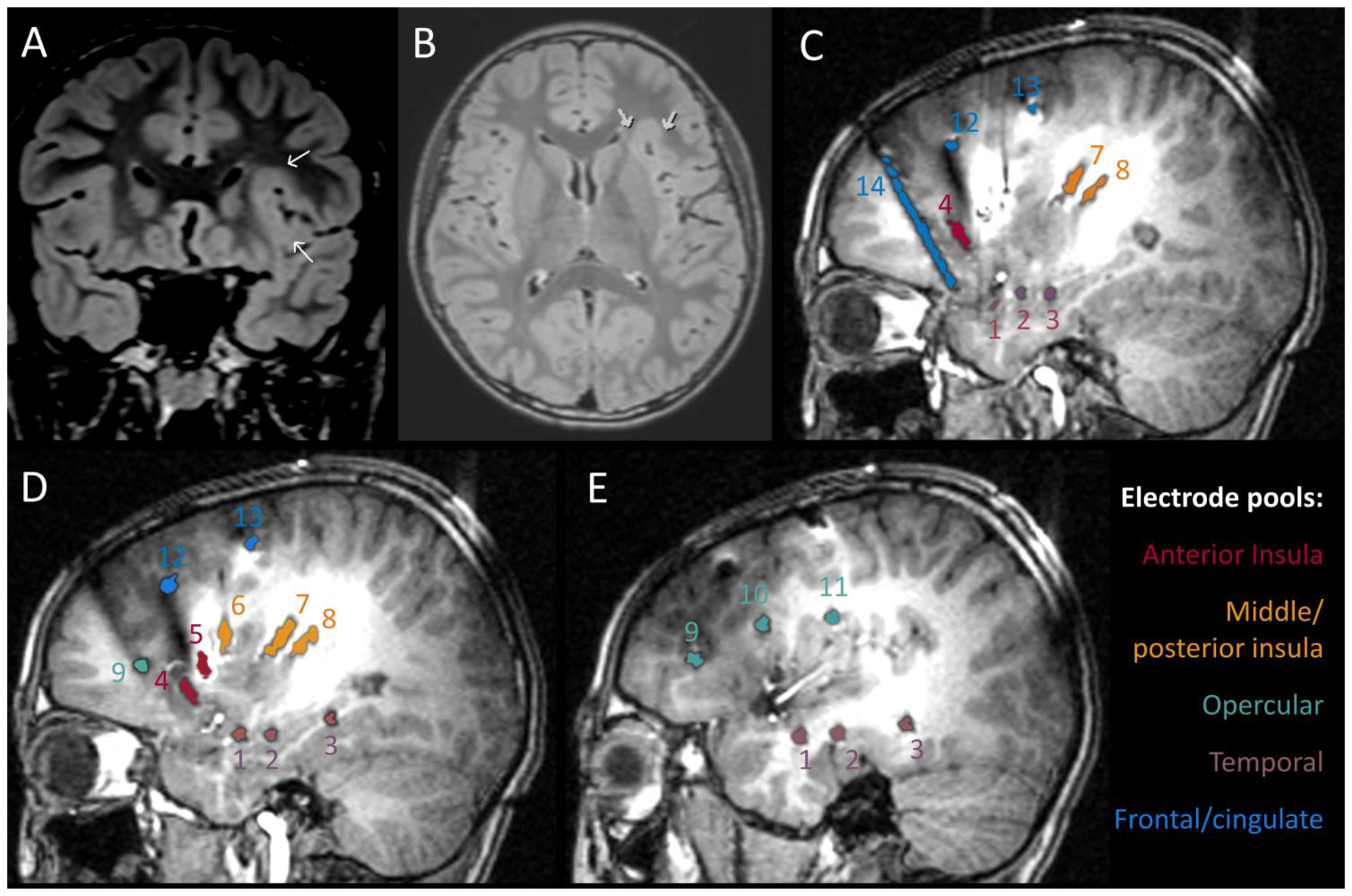
Magnetic resonance imaging showing focal cortical dysplasia and electrode pools. Coronal (A) and axial (B) FLAIR sequence showing increased cortical thickness and blurred gray-white matter junction in the left anterior insula (arrows). C-E: Sagittal T1 sequences from left medial (C) to lateral (E) showing the localization and pooling (different colors) of the electrodes. 1: amygdala, 2: anterior hippocampus, 3: posterior hippocampus, 4: anterior insula – anterior, 5: anterior insula – posterior, 6: middle insula, 7: posterior insula – anterior, 8: posterior insula – posterior, 9: frontal operculum – anterior, 10: frontal operculum – posterior, 11: central operculum, 12: anterior cingulate, 13: middle cingulate, 14: lateral orbitofrontal

He underwent sEEG using 14 depth electrodes in the left hemisphere to delineate the extent of the seizure onset zone (Fig. 1 C-E). Frequent interictal epileptiform discharges were recorded arising from the left anterior insula, at times with a field involving the left middle and posterior insula as well as the left lateral orbitofrontal region. Less frequently, interictal epileptiform discharges were seen arising from the left mesiotemporal and lateral temporal region. Twenty-two seizures (20 electroclinical, 2 electrographic) were captured arising from the left anterior insula with propagation to the left middle insula and sometimes further spread to the left posterior insula, frontal and mesiotemporal electrodes (Fig. 2). Based on the results of the recording, the patient underwent LITT surgery of the left anterior insula with significant improvement of his seizure frequency 5 months postoperatively (mild focal aware seizures every few weeks).

**Figure 2:**
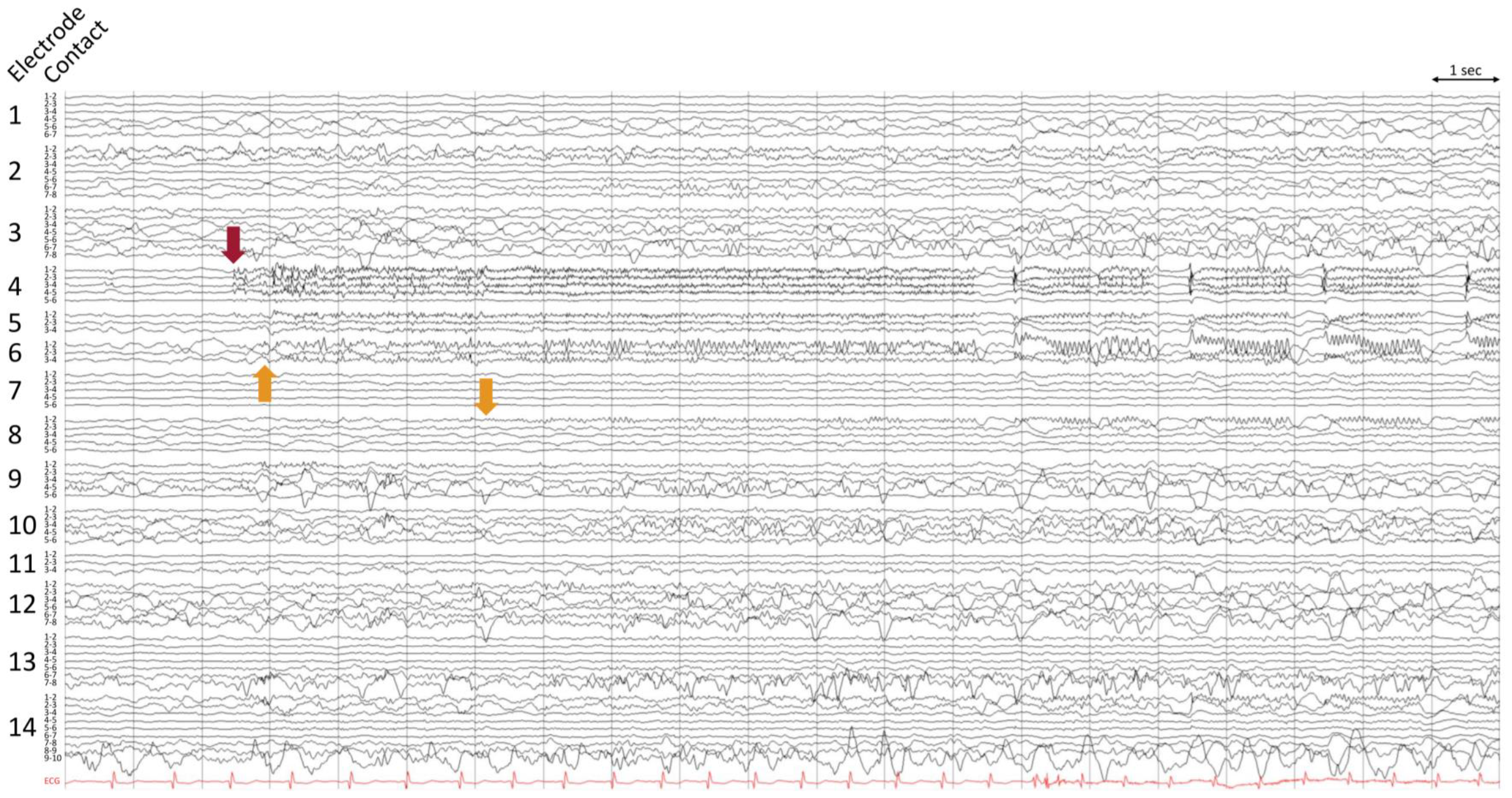
EEG onset in left anterior insula. Stereo EEG recording of a typical electroclinical seizure (low frequency filter 1 Hz, high frequency filter and notch off, sensitivity 100 μV/mm, 30 mm/sec). The red arrow indicates seizure onset at electrode 4 (left anterior insula – anterior) with lower amplitude involvement of the deep contacts of electrode 5 (left anterior insula – posterior). Yellow arrows: Propagation to electrode 6 (middle insula) after ∼0.5 sec and to the deep contact of electrode 8 (left posterior insula – posterior) after ∼3.5 sec.

The electrodes were combined into 5 pools for genetic analysis (Fig 1 C-E, Table 1). Due to the presence of the lesion and seizure onset zone in the left anterior insula, the left anterior insula pool was selected as the affected region. The left frontal/cingulate pool was selected as unaffected control region as it was distantly located from the affected pool and no epileptiform activity was seen arising primarily from this area.

**Table 1:**
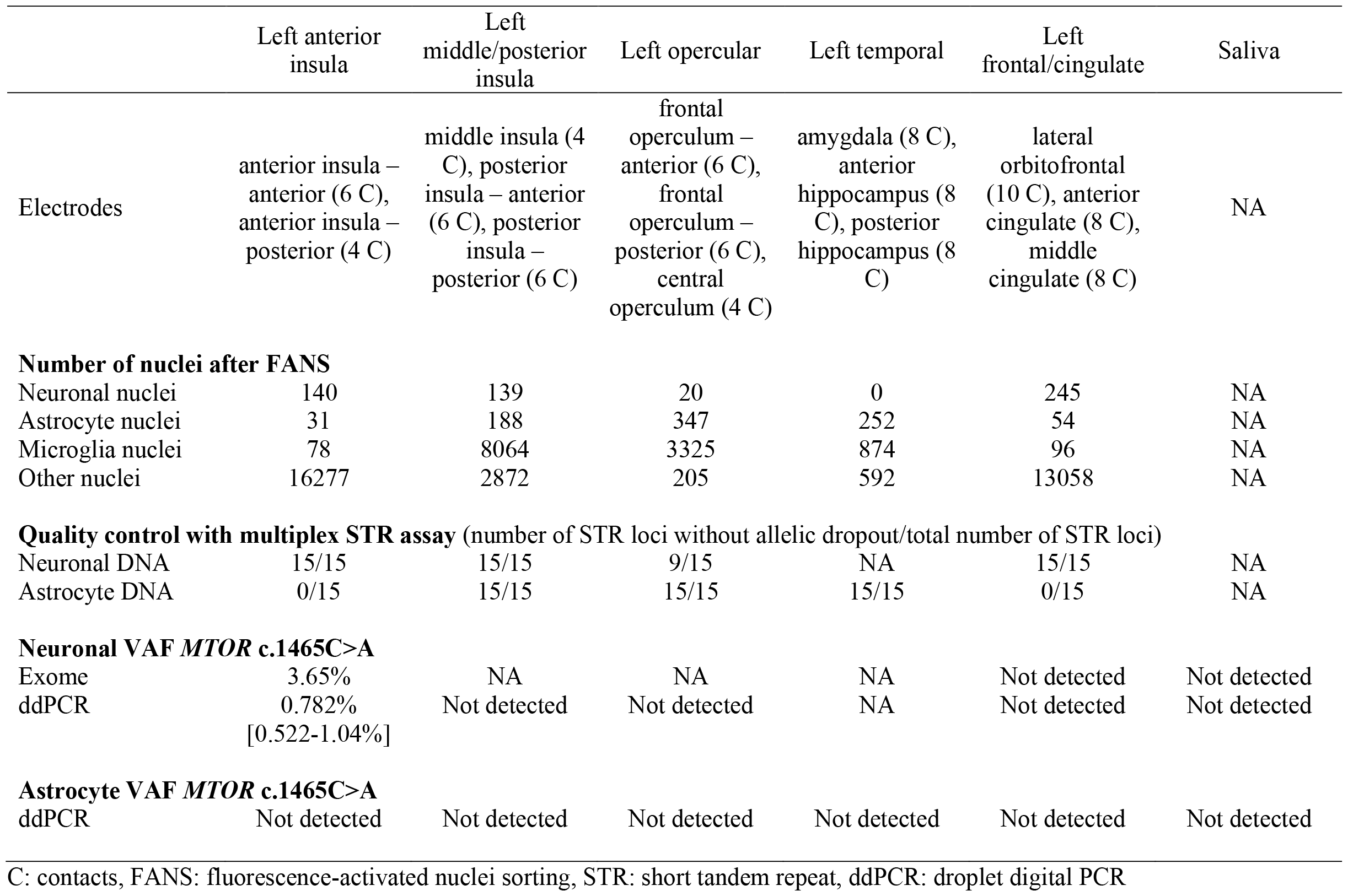
Depth electrode pools with nuclei yield and variant allele frequencies.

FANS isolated 140 neuronal nuclei from the affected depth electrode pool and 245 neuronal nuclei from the unaffected depth electrode pool (Table 1). DNA from neuronal and astrocyte nuclei of all pools was amplified using PTA. The multiplex assay revealed identical alleles for all 15 STR and the one sex marker in both neuronal samples and saliva excluding contamination and confirming excellent amplification (Table 1).

Exome sequencing resulted in a mean coverage of 178x in the affected neuronal sample with 79% of the exome covered at ≥95x. The unaffected neuronal sample had a mean coverage of 258x with 91% of the exome covered at ≥95x. A novel mosaic missense *MTOR* variant (ENST00000361445:c.1465C>A, p.Leu489Met) was identified in the affected neuronal sample at a VAF of 3.65% (or 6 reads) which was not present in the unaffected neuronal sample and saliva. The variant had a CADD v1.6 score of 23.7^34^, REVEL score of 0.632^35^, and MISTIC score of 0.863^36^. The variant was not present in gnomAD^31^ and TOPmed^32^. Droplet digital PCR confirmed the mosaic gradient with a VAF of 0.78% in the affected neuronal sample (Fig. 3, Table 1). The variant was not present in any other neuronal or astrocyte sample. The detection limit of the ddPCR assay was 0.05% VAF.

**Figure 3:**
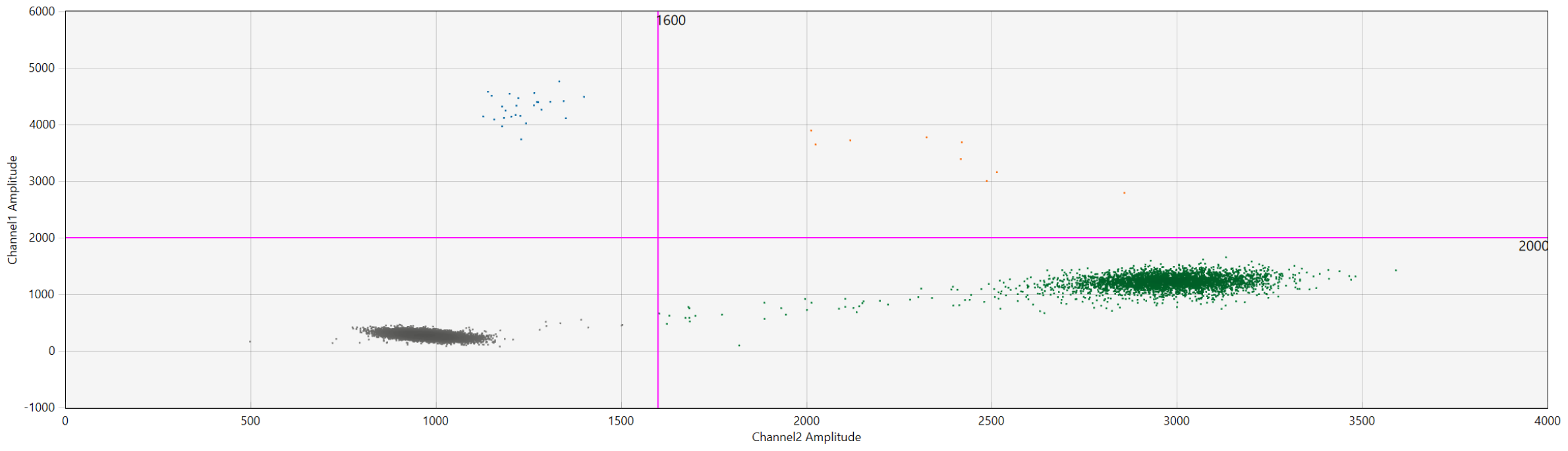
Droplet digital PCR result for left anterior insula pool. Gating was determined based on fluorescent intensities of no template control samples. Variant concentration: 2.75 copies/μL, wild-type concentration: 348 copies/μl, variant allele fraction: 0.782%. Blue: variant droplets, green: wild-type droplets, orange: droplets containing multiple DNA templates, gray: empty droplets.

## Discussion

We achieved molecular diagnosis of a novel somatic mosaic *MTOR* missense variant in a patient with genetic-structural focal epilepsy due to left anterior insula FCD using purified neuronal DNA harvested from sEEG depth electrodes. Mosaic missense variants in *MTOR* have been reported in the past in FCD type II indicating perfect gene-disease validity^2–9,11^. Furthermore, the variant is not seen in control cohorts and the high CADD score predicts deleteriousness. Additional support for the pathogenicity of the variant is provided by the mosaic gradient with the highest VAF in the lesion and seizure onset zone and absence of the variant in other brain regions and saliva. Assuming even amplification, a VAF of 0.78% corresponds to 1.56% of the neurons in the affected pool carrying the variant, i.e. 2 out of 140 neurons.

Our finding confirms that harvesting neuronal DNA from depth electrodes followed by molecular analysis to identify brain somatic variants is feasible. We achieved molecular diagnosis in the absence of resected surgical tissue as the patient underwent LITT surgery so no tissue could be harvested. In our patient, harvesting cells adhering to depth electrodes was the only available access to brain DNA. As minimally invasive surgical approaches are increasingly being used, it is of paramount importance to further develop techniques to identify somatic variants independent of the availability of surgically resected tissue. With the continuing development of precision medicine approaches, for example the use of mTOR inhibitors in mTORopathies^37,38^, such techniques will be paramount to allow optimal treatment of patients in the future.

We posit that the success in our patient was made possible by our novel method that allows us to investigate pure neuronal DNA by using FANS to exclude non-brain cells. This method has multiple advantages.

1. Our technique provides the exact number of nuclei of each cell type that are being amplified. This allows estimating the power to identify a certain VAF. In our case, the variant was not seen in the left middle/posterior insula pool which was located most closely to the affected anterior insula region. The presence of 139 neuronal and 188 astrocyte nuclei in this pool provided 80% power to identify VAFs of 0.6% and 0.5%, respectively. We can also conclude that the number of isolated neuronal or astrocyte nuclei of a few other regions was insufficient to exclude low percentage mosaicism (left opercular neuronal nuclei, n=20; left anterior insula astrocyte nuclei, n=31; left frontal/cingulate astrocyte nuclei n=54). This was also reflected by low quality on the STR multiplex assay for these samples whereas all other samples showed excellent quality (Table 1). No neuronal nuclei were isolated from the left temporal pool and, hence, no neuronal DNA was available from this region.
2. The exclusion of non-brain cells by FANS increases the frequency of neuronal variants resulting in a higher chance to identify low percentage mosaicism (low VAF). The previous depth electrode harvesting method amplified DNA from the bulk sample. Any contamination of the sample, for example by blood or immune cells, reduced the percentage of brain cells and resulted in lower VAFs that are more difficult to detect.
3. Our method allows processing of all cells of an electrode pool and then purifies the cell type of interest. Previous methods used only 8% (4 out of 50 μl)^16^ or 30% (3 out of 10 μl)^20^ of the sample which avoids exceeding the maximum number of cells that can be processed with the amplification kit. However, this means that only a subset of the neurons in a pool were amplified. If fewer neurons undergo amplification, there is a higher risk of missing the abnormal cell fraction in the sample which may prevent detection of the variant with the previous method.

Depth electrode harvesting methods in general are limited by the low quality of the cells that are harvested from depth electrode samples. Using our method, a large percentage of nuclei (97%) did not stain as neuronal, astrocyte or microglia. One explanation for this finding is that these nuclei are indeed not of brain origin and represent contamination. In this case, it will be very important to exclude these cells before amplification due to the aforementioned reasons. However, it is possible that some of the unstained nuclei may originate from cells of brain origin but due to degradation they do not stain properly. As successful whole genome amplification requires high molecular DNA, it is likely that amplification from degraded cells with fragmented DNA will not be successful and, hence, exclusion of this pool would also be beneficial.

The observed VAF (0.78%; representing only 2 of 140 neurons in the pool) is within the range of *MTOR* VAFs previously reported in FCD^6,39^. However, it is likely that the VAF at the SOZ was somewhat underestimated using this approach due to the dilution of the sample with healthy cells from other areas that were penetrated by the electrodes as well.

In this study, the variant was only identified in the neuronal nuclei, supporting the concept that the variant was present in the expected cell type for the disease. However, as only 31 astrocyte nuclei were isolated from the affected pool, it is possible that a low percentage of astrocytes carrying the variant were missed. As future research investigates the presence of mosaic variants in seizure foci, we expect that some identified variants may not be restricted to the neuronal cell type, depending on when the mosaic variant occurred in embryogenesis or neurodevelopment^13^. Non-neuronal cells have been implicated in epileptogenesis^40–42^; the possibility of somatic variants in non-neuronal cells associated with epilepsy is conceptually intriguing and could be detected using this approach.

In summary, we used our novel depth electrode harvesting method to establish a molecular diagnosis of a somatic mosaic *MTOR* variant in an adolescent with genetic-structural focal epilepsy due to left anterior insula FCD. Our novel method represents a significant improvement compared to the previous depth electrode harvesting method by focusing the analysis on high quality cells of the cell type of interest. It will pave the way for molecular diagnosis in patients who do not undergo resective epilepsy surgery and allow application of precision medicine approaches based on the genetic finding.

## Data Availability

The de-identified phenotypic and exome sequencing data will be made available upon reasonable request from any qualified investigator after institution of a material transfer agreement.

## Acknowledgements

We would like to thank the patient and his family for participation in the study. We acknowledge the Flow Cytometry Core Facility and the Centre for Health Genomics and Informatics at the University of Calgary for their support.

## Author contributions

Karl Martin Klein: Designed and conceptualized the study; major role in the acquisition of data; analyzed the data; drafted the manuscript for intellectual content; supervision; funding acquisition.

Rumika Mascarenhas: Major role in the acquisition of data; analyzed the data; reviewed the manuscript for intellectual content

Daria Merrikh: Major role in the acquisition of data; reviewed the manuscript for intellectual content

Maryam Khanbabaei: Major role in the acquisition of data; reviewed the manuscript for intellectual content

Tatiana Maroilley: Analyzed the data; reviewed the manuscript for intellectual content Navprabhjot Kaur: Major role in the acquisition of data; reviewed the manuscript for intellectual content

Yiping Liu: Major role in the acquisition of data; analyzed the data; reviewed the manuscript for intellectual content

Tyler Soule: Major role in the acquisition of data; reviewed the manuscript for intellectual content

Minette Manalo: Major role in the acquisition of data; reviewed the manuscript for intellectual content

Goichiro Tamura: Major role in the acquisition of data; reviewed the manuscript for intellectual content

Julia Jacobs: Major role in the acquisition of data; reviewed the manuscript for intellectual content

Walter Hader: Major role in the acquisition of data; reviewed the manuscript for intellectual content

Gerald Pfeffer: Analyzed the data; reviewed the manuscript for intellectual content Maja Tarailo-Graovac: Designed and conceptualized the study; analyzed the data; drafted the manuscript for intellectual content; supervision; funding acquisition

